# Exploratory autoantibody profiling in autism spectrum disorder

**DOI:** 10.1101/2025.10.06.25337143

**Authors:** Melody P. Lun, Thomas T. Ngo, Iris Tilton, Ravi Dandekar, Margaret Pekar, Audrey Thurm, Christopher M. Bartley, Michael R. Wilson, Samuel J. Pleasure

## Abstract

There is growing evidence that immune dysfunction interacts with genetic predisposition to increase vulnerability to autism spectrum disorders (ASD). However, few studies have extensively profiled the autoantibody repertoire from children with ASD. Here, we utilized unbiased approaches to identify the antigenic targets of autoantibodies from cerebrospinal fluid (CSF) and blood collected from children with ASD compared to typically developing controls. In a cohort of children with ASD, we identified 6 of 61 participants (9.8%) harbored anti-neural autoantibodies in their CSF with distinct immunoreactivity patterns by tissue-based immunofluorescence screening on murine brain sections. In one participant, two CSF samples collected 2.3 years apart showed persistent anti-neural immunofluorescence. Phage display immunoprecipitation sequencing (PhIP-seq) and immunoprecipitation mass spectrometry (IP-MS) were utilized to screen for the antigenic targets of these CSF immunoreactivities, which revealed that each of these 6 cases have unique autoreactivities in their CSF as well as their peripheral blood. Our screening techniques identified several candidate autoantigens that have strong genetic associations with ASD, including ANK2/3, BCL11A, CHD3/8, NRXN1/2, RUNX1T1, and ZNF292. Broadly, these candidate autoantigens participate in several pathways that may contribute to ASD, including synaptic connectivity, neuronal scaffolding, and transcriptional regulation. While the autoantibody specificities remain to be validated through orthogonal assays, these data demonstrate the potential for parallel unbiased screens to identify autoantibodies with relevance to ASD.

## INTRODUCTION

ASD is a neurodevelopmental disorder characterized by persistent and pervasive social communication deficits and restricted and repetitive behaviors^1^. ASD typically presents early in childhood, although the progression and spectrum of clinical features is heterogeneous and often accompanied by other psychiatric comorbidities^2^. The prevalence of ASD has steadily increased in recent decades^3^, with recent estimates that 1 in 31 (3.2%) of school-aged children in the United States are affected^4^. The genetic architecture of ASD has been partially mapped through whole-genome sequencing of large ASD cohorts, revealing that genetic variants account for ∼14% of individuals with ASD^5^. However, the majority of cases remain unexplained, suggesting that environmental and immune factors may interact with genetic susceptibilities to contribute to the neurobiological and behavioral abnormalities in ASD.

From early fetal development through adulthood, there is an essential interplay between the immune and central nervous systems (CNS)^6,7^. Both innate and adaptive immunity regulate brain development, from neuronal migration to synaptic pruning and circuit assembly. In the earliest stages of brain development *in utero*, maternal immune dysfunction secondary to infectious and inflammatory events in pregnancy likely underlies a subset of ASD^8^. This linkage between maternal infections and increased risk of ASD was identified following the rubella epidemic in the United States in 1964^9^. Thereafter, several large population registry studies have also identified an overrepresentation of parental history of autoimmunity among children with ASD^10–12^. Autoantibodies from mothers of children with ASD have been shown to react against fetal brain proteins^13,14^. Several antigenic targets of maternal autoantibodies linked to ASD have been identified^13,15–17^, but whether these autoantibodies are pathogenic remains to be demonstrated.

As brain development continues past infancy, immune dysregulation during early childhood may similarly contribute to risk of ASD. Since ASD is heterogeneous with a wide spectrum of severity, it has been proposed that autoimmune encephalitis, or inflammation of the brain secondary to an individual’s immune dysfunction, may be a causative event leading to an acquired form of ASD or may produce symptoms misdiagnosed as ASD^18^. Neuroinflammation contributing to a subset of acquired ASD suggests the use of immune-modulating agents as possible treatments for ASD symptoms, though a trial of intravenous immunoglobulins (IVIG) in ASD had mixed results^19^, and evidence of its benefit through rigorous randomized controlled trials is lacking. Nevertheless, identification of specific autoantibodies as potential biomarkers may enable improved precision in diagnosis and expedite early behavioral intervention.

Few studies of ASD have interrogated the immune repertoire from the CNS, instead focusing on the peripheral blood. A prior publication, based on the same longitudinal cohort of children with ASD studied here, had assessed immune mediators in paired CSF and peripheral blood but did not identify significant differences in the expression of circulating cytokines and chemokines between children with ASD versus typically developing controls, suggesting that active inflammatory processes are unlikely to play a role in the maintenance of ASD^20^. Moreover, among participants with longitudinally collected samples, there was high within-subject variability in concentrations of analytes across time points, likely reflective of the immune milieu in the acute period. While chemokines and cytokines mirror the immediate inflammatory state, autoantibodies provide an index of longer standing, sustained immune states. While there have been prior attempts to profile autoantibodies in peripheral blood, the findings have been inconsistent across studies with some lacking identification of an autoantigen^19,21–23^. To our knowledge, there are no prior studies that have investigated the CSF autoantibodies in ASD. The goal of this study was to profile the CSF autoantibodies from children with ASD since investigation of the CNS immune milieu may yield relevant autoantibodies that were not previously identified peripherally in blood.

## MATERIALS AND METHODS

### Participants and sample collection

Recruitment for a longitudinal study of ASD has been described previously^20,24,25^. Participants for the current study were drawn from a longitudinal study of ASD (NCT00298246) implemented at the National Institute of Mental Health, Bethesda, Maryland. Eligible participants met DSM-IV-TR criteria for ASD on the basis of the Autism Diagnosis Interview-Revised (or a Toddler version)^26^ and the Autism Diagnostic Observation Schedule (ADOS)^27^. Written informed consent was obtained from the guardians of each participant. The longitudinal study included 106 children with ASD; 67 of these participants underwent at least one lumbar puncture (LP) for research purposes during the study period of 2006-2013 and were included in this current study. A subset of these children (n = 31) had a second LP. Exclusion criteria included neurologic conditions such as cerebral palsy, or aggressive or maladaptive behaviors that precluded participation in behavioral testing. Fifty typically developing children were included as controls, screened by the Social Communication Questionnaire^28^ and cognitive testing without concern in any developmental domain. The groups were not matched on cognitive function. All children in the current study were between the ages of 2 and 7.99 years at baseline and were followed at intervals of approximately one year, for up to 3 years. Per study design, blood samples were collected from subjects up to four times at intervals to approximately 1 year, and CSF samples were collected once (at baseline) or twice (at final visit, if possible).

Medical history related to immune status (i.e., allergies, immunodeficiency, autoimmune disorder) was collected for both groups of children via structured interview with medical personnel. For subjects in the ASD group who underwent LP, serum was collected contemporaneously with CSF, under sedation, following a 12-hour fasting period. All CSF and blood samples were collected during in the morning (9am–12pm). Ethical constraints prevented LPs in the control group. Control serum samples were not collected under sedation. CSF was immediately centrifuged after the LP procedure, and acellular aliquots were stored at −70°C within 20–30 minutes. Serum samples were collected following standard procedures, aliquoted, and stored within 20–30 minutes after separation. Up to four serum samples and up to two CSF samples were obtained from each participant at intervals ranging from 9–24 months and stored until simultaneous laboratory analysis.

### Animal use

C57BL/6J mice (Jackson Laboratory) were bred and collected on postnatal day (P)28, when murine neurodevelopment approximates toddlerhood and early childhood in humans. P28 brain tissue was used for immunostaining, immunoprecipitations (IPs), and immunoblotting. All animal procedures complied with policies of the UCSF Institutional Animal Care and Use Committee (AN204753).

### Mouse brain tissue-based immunofluorescence

P28 mice were perfused with ice-cold PBS, followed by 4% paraformaldehyde (PFA), and post-fixed overnight. Brains were embedded and cryosectioned sagittally at 12μm thickness. Sections were permeabilized and blocked with 10% lamb serum in 0.1% Triton-X 100 in PBS, incubated with CSF diluted 1:4 in blocking solution at 4°C overnight, followed by Alexa Fluor-conjugated secondary antibodies (Jackson ImmunoResearch 709-545-149). To identify co-localization of patient CSF autoantibodies and candidate autoantigens, sections were incubated with CSF together with the following commercially available antibodies: rabbit anti-NRNX2 (Proteintech 30490-1-AP).

### Programmable phage display immunoprecipitation sequencing (PhIP-seq)

We adapted a previously published protocol for PhIP-seq^29,30^, which utilizes a human peptide library of 731,724 unique peptides that are displayed by T7 bacteriophage, with each phage expressing a different 49-amino-acid peptide on its surface. In total, these peptides tile the entire human proteome, including all known isoforms, with an overlap of 25 amino acids between neighboring peptides. CSF and serum samples were incubated with 10^10^ plaque forming units per mL of library, and antibody-bound phage were captured with protein A/G magnetic beads (ThermoFisher 10001D, 10003D). The beads were washed and used to inoculate *Escherichia coli* (BLT5403, EMD Millipore) to amplify phage for a second round of IP with the same patient’s CSF or serum. DNA was collected by phage lysis, PCR amplified and barcoded with TruSeq dual indices. PCR products were pooled and bead cleaned (SPRISelect, Beckman Coulter), and DNA quantity and integrity were analyzed by Tapestation (Agilent Technologies). Pooled libraries were sequenced on Illumina NovaSeqX using 150bp paired-end reads with a goal average read depth of 2 million reads per sample.

### PhIP-seq analysis

Data analysis was performed using the R programming language. Raw FASTQ reads were trimmed and aligned to the reference PhIP-seq database using RAPSearch (v2.2). Peptide counts were normalized to reads per 100,000 (RPK) for each sample by dividing each peptide count by the sum and multiplying by 100,000. The resulting peptide RPK counts were normalized against bead-only mock IPs, and these bead-normalized peptide counts were used for all subsequent analyses.

For analysis of serum samples, data were divided into ASD and control groups. For analysis of CSF from children with ASD, samples with positive tissue-based immunofluorescence (“Case_Pos”) were compared against samples with negative tissue-based immunofluorescence (“Case_Neg”). For analysis of serum, samples from children with ASD (“Case”) were compared against samples from typically developing children (“Control”).

A peptide was determined to be enriched if the bead normalized RPK ≥ 10 and Z-score value ≥ 3 relative to controls across both technical replicates. To calculate Z-scores, controls for CSF were negative-staining samples from children with ASD (all “Case_Neg” samples), whereas controls for serum were from typically developing children. At the peptide level, a Wilcoxon rank-sum test with a Benjamini-Hochberg (BH) adjustment was used to determine statistical significance of the bead-normalized peptide counts between cases and controls (adjusted P-value < 0.05).

We identified peptide enrichment shared across multiple CSF or serum samples from “cases” by 1) comparing all positive-staining CSF samples (“Case_Pos” samples) versus all negative-staining CSF samples (“Case_Neg” samples) and 2) comparing all serum samples from children with ASD (“cases”) versus all serum samples from typically developing children (“controls”). At this group level analysis for CSF and serum, peptides were enriched if they met these additional filters: Number of cases meeting enrichment criteria (bead normalized RPK ≥ 10 and Z-score value ≥ 3) in both technical replicates ≥ 2 for CSF and ≥ 5 for serum, sum RPK of a given peptide fragment across all cases > 100, case to control sum RPK ratio of a given peptide fragment > 2, and case to control sum bead-normalized RPK ratio of a given peptide fragment > 2.

We also identified peptide enrichment for each CSF with positive staining (“Case_Pos” samples) at the individual participant level by identifying the most enriched peptides in the individual above other cases. Case to control sum RPK ratios were adjusted to identify the most enriched peptides for an individual’s case-specific heatmap. Peptides from large proteins, including TTN and OBSCN, were omitted from further analysis.

### Immunoprecipitation-Mass Spectrometry (IP-MS)

Whole mouse brain lysates were prepared from P28 male and female C57BL/6J mice. One male and one female were pooled for each technical replicate. Brains were rapidly dissected into ice-cold PBS, homogenized with a dounce homogenizer in lysis buffer (1% Triton-X 100 in TBS with 2X HALT Protease and Phosphatase Inhibitors, ThermoFisher). Brain lysates were centrifuged at 4°C for 10 minutes at 10,000x*g*. The supernatant was collected and diluted to 5μg/μL. IPs were performed in technical replicate using patient CSF. 25μL of CSF was added to 200μL of mouse brain lysate in Protein Lo-bind tubes (Eppendorf), incubated for 90 minutes with rotation at 4°C followed by 90 minutes with rotation at room temperature. 10μL of 1:1 mixture of magnetic protein A/G beads (ThermoFisher 10001D, 10003D) were added to each sample and incubated for 1 hour at 4°C. Beads and immune complexes were captured on a magnetic stand and washed three times with detergent wash buffer, a high-salt buffer, a non-detergent wash buffer, and finally with ammonium bicarbonate buffer. All wash buffer was removed, and beads were stored at -80°C.

Beads were submitted to UC Davis Proteomics Core for digestion and LC-MS/MS. Beads were washed in ammonium bicarbonate buffer, and proteins were digested with trypsin overnight at room temperature. After digestion, the supernatant was removed and saved, the beads were washed with ammonium bicarbonate, and the supernatant was removed and saved with the previous extract.

NanoLC was performed on an Evosep One instrument (Evosep Biosystems, Denmark) and eluted directly onto a PepSep analytical column. MS was performed on a hybrid trapped ion mobility spectrometry-quadrupole time of flight mass spectrometer (timsTOF HT, Bruker Daltonics, Bremen, Germany) with a modified nano-electrospray ion source (CaptiveSpray, Bruker Daltonics) in PASEF mode. Desolvated ions entered the vacuum region through the glass capillary and deflected into the TIMS tunnel which is electrically separated into two parts (dual TIMS). The dual TIMS analyzer was operated at a fixed duty cycle close to 100% using equal accumulation and ramp times of 85 ms each. The data-independent analysis (DIA) scheme consisted of one MS scan followed by MS/MS scans taken with 36 precursor windows at width of 25Th per 1.09 sec cycle, over the mass range 300-1200 Dalton. DIA data were analyzed using Spectronaut 18.4 (Biognosys Schlieren, Switzerland) using the direct DIA workflow with PTM localization selected. Trypsin/P specific was set for the enzyme allowing two missed cleavages. Fixed modifications were set for carbamidomethyl, and variable modifications were set to acetyl (Protein N-term), and oxidation.

For each IP, a fold change of protein quantity in the IP with positive-staining case CSF was compared to negative-staining CSF from Case 3 and 4 and bead-only IP controls. Statistical analysis was performed by Student’s t-test with BH correction. For each case, proteins were filtered by log2(fold change) > 1 and FDR-adjusted p value < 0.05. All proteins that met these criteria were considered “present” in the IP.

Proteins present by IP-MS were overlapped with the PhIP-seq enriched peptides for each case. Transcripts per Million (TPM) data from the Human Protein Atlas (HPA) Brain Resource^31^ (downloaded on September 10, 2025) were used and overlapped with the gene labels of enriched peptides. The HPA brain expression data (cerebral cortex, hippocampal formation, and cerebellum) were used in all case-specific heatmaps. A log10 transform was applied to all plots. To filter for brain-enriched genes, a threshold of sum log10(TPM) > 2 was used for the select overlapping HPA data. For Case 2, the threshold was increased to sum log10(TPM) > 3.

### Cell-based overexpression assay

HEK293T cells (American Type Culture Collection CRL-3216) were plated onto 10mm coverslips coated with poly-d-lysine. Cells were transfected with the following FLAG- or HA-tagged expression constructs using Lipofectamine 3000 (ThermoFisher L3000015): NRXN1 (Addgene #58266), NRXN2 (Genscript OHu10454), and empty pcDNA3.1+ c-DYK vector (Genscript). After 24 hours, cells were washed with ice-cold PBS and fixed with 4% PFA for 10 minutes. Cells were permeabilized, blocked, and immunostained overnight with mouse anti-HA antibody (Proteintech 66006-2-Ig at 1:500), mouse anti-FLAG antibody (Millipore F3165 at 1:2000) and patient CSF at 1:10 dilution or patient serum at 1:200 dilution followed by Alexa Fluor-conjugated secondary antibodies (Jackson ImmunoResearch 715-585-151, 709-545-149).

### Microscopy

Stitched composite images of sagittal mouse brain sections were acquired on Zeiss AxioScan.Z1 at 20x magnification. Confocal images of mouse brain sections and HEK293T overexpression cell-based assays were acquired at 40X at the UCSF Nikon Imaging Center using a Nikon CSU-W1 spinning disk confocal microscope.

### IP and Western blot

Cell lysates from HEK293T transfected cells were collected and immunoprecipitated with anti-HA (ThermoFisher 88836) or anti-FLAG coated magnetic beads (MedChemExpress HY-K0207). Beads were washed, and immunoprecipitated protein was eluted from the beads. Protein was loaded into the wells of a 4-12% Criterion XT Bis-Tris Protein Gel (Bio-Rad), separated at 180V for 1 hour, and transferred to polyvinylidene fluoride (PVDF) membranes (Bio-Rad) at 100V for 1 hour on ice. PVDF membranes were blocked with Intercept PBS Blocking Buffer (LICORBio 927-60001), incubated with CSF diluted 1:10 in blocking buffer overnight at 4°C, followed by goat anti-human IRDye 800CW (LICORbio 926-32232), anti-HA conjugated to DyLight800 (Invitrogen 26183-D800), or anti-FLAG conjugated to Dylight680 (Rockland 200-344-383). Blots were imaged on Li-Cor Odyssey.

## RESULTS

### Detection of anti-neural antibodies in ASD CSF by tissue-based immunofluorescence

84 CSF samples from 61 children who met ADOS criteria for ASD were screened for presence of autoantibodies targeting neural antigens by immunofluorescence on mouse brain tissue. CSF from 6 out of 61 children (9.8% of ASD participants) exhibited anti-neural immunoreactivity (**Figure 1A**). Of these 6 children, 3 had a follow-up LP but only one (Case 2) had anti-neural autoantibodies in both CSF samples collected over a 2.3 year period, whereas two children (Case 3 and 4) had anti-neural autoreactivity in the follow-up CSF, but negative anti-neural autoreactivity on the initial CSF. Each case demonstrated a distinct anatomical immunofluorescence pattern with unique subcellular localization (**Figure 1B-C**), including nuclear, cytoplasmic, and neuropil staining, suggesting that the antigenic targets of CSF autoantibodies differ in each case.

**Figure 1.**
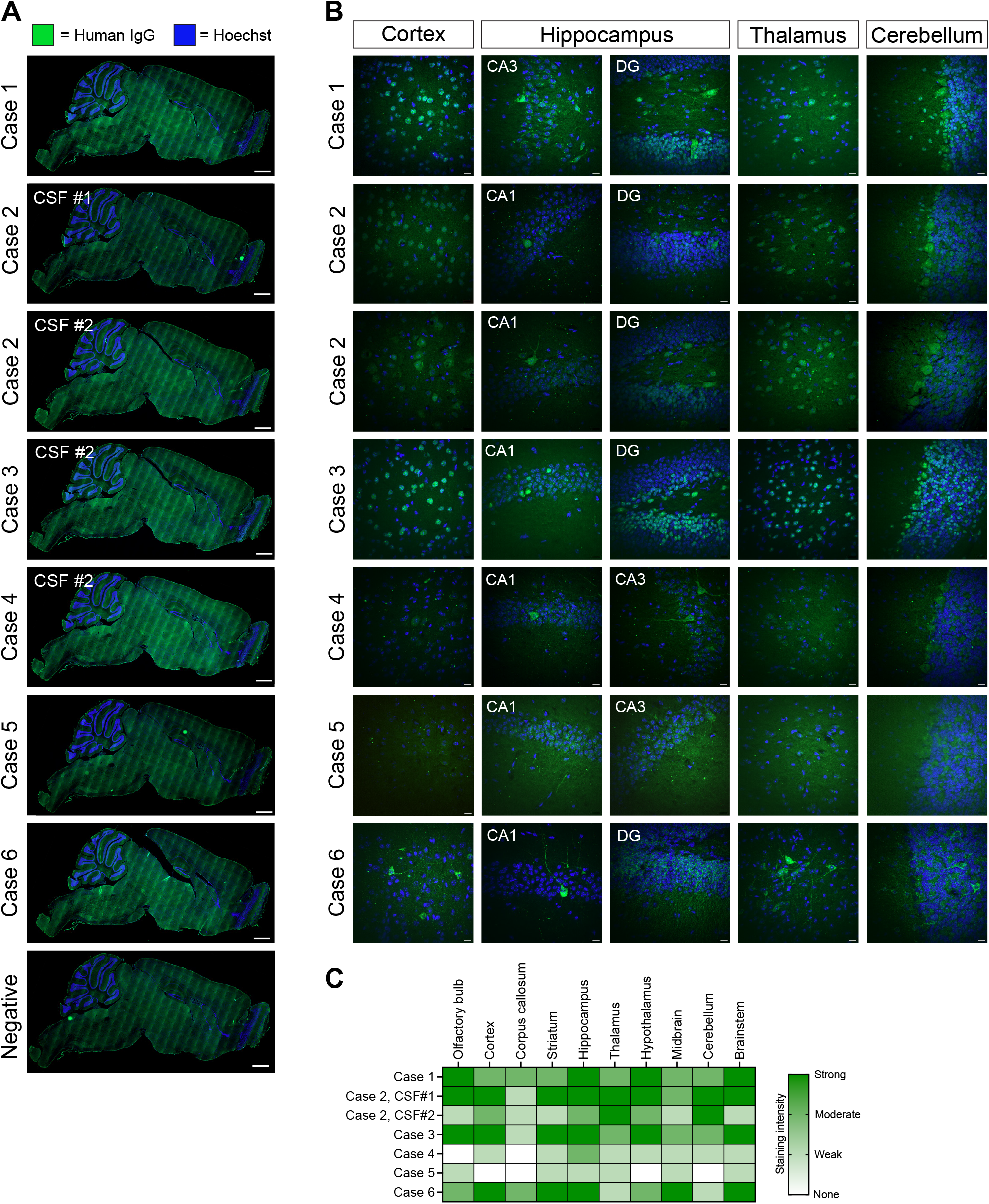
Detection of anti-neural antibodies in CSF of children with ASD. **(A)** Sagittal P28 mouse brain sections were immunostained with CSF at 1:4 dilution (human IgG, green). Anti-human IgG-488 secondary only negative control was not immunoreactive, compared to 7 CSF samples from 6 ASD children (Cases 1-6). Scalebars, 1000μm. **(B)** Select high-magnification confocal images of immunostained mouse brain sections from cortex, hippocampus (CA1, CA3, dentate gyrus), thalamus, and cerebellum. Scalebars, 20μm. **(C)** Heatmap indicating presence and strength of immunoreactivity signal across immunostained mouse brain sections.

The clinical features of each case were variable, though by parental report, all 6 cases were noted to have developmental delays in infancy without acute regression of developmental milestones (**Table 1**). Compared to other ASD participants, these 6 cases did not demonstrate significant abnormalities in basic CSF composition, including white blood cell counts, glucose or protein levels. The IgG index across all cases were within normal range, although notably the albumin quotient for Case 2 was elevated in both CSF samples, suggesting increased blood-brain barrier permeability.

**Table 1.** Participant demographic and phenotypic information.

|  | Summary for all ASD participants | Case 1 | Case 2 | Case 3 | Case 4 | Case 5 | Case 6 |
| --- | --- | --- | --- | --- | --- | --- | --- |
| Sex | 82% male | Male | Female | Female | Male | Male | Male |
| Age at ASD diagnosis (yrs) | 3.60 ± 0.95 | 3-5 years | < 3 years |  | 3-5 years | < 3years | 3-5 years |
| Medical history |  |  | Epilepsy |  | Congenital syphilis, Recurrent otitis media, Reactive airway disease | Twin brother with ASD, Neonatal sepsis | Prematurity (late preterm) |
| Brain MRI |  |  | Normal |  | White matter signal hyperintensities in bilateral cerebellar hemispheres and left anterior hypothalamus, resolved on subsequent imaging | Normal | Two foci of abnormality in right frontal lobe subcortical white matter |
| Genetics |  |  | Unbalanced translocation (single copy gain of 2p25.3 and single copy loss of 9p24.3) |  | Unremarkable, Normal | Unbalanced translocation in chr 2, 8, and 9 |  |
| Developmental milestones |  |  | Hypotonia, delayed walking |  | Hypotonia, speech and motor delays | Severe communication disorder | Expressive language delay, Poor eye contact and arm flapping |
| **Behavioral characteristics at baseline** |  |  |  |  |  |  |  |
| Vineland II composite standard score |  |  | 65 |  | 68 |  |  |
| ADOS calibrated severity score | 7.71 ± 1.6 |  | 6 |  | 6 |  |  |
| **CSF composition** |  |  |  |  |  |  |  |
| CSF WBC count, 0-5/mm3 (mean ± SD) | 0.90 ± 1.02 | N/A | CSF#1: 5<br>CSF #2: 5 | CSF #1: 0<br>CSF #2: 1 | CSF #1: 1<br>CSF#2: 1 | 0 | 0 |
| CSF glucose, 40-70 mg/dL (mean ± SD) | 59.82 ± 5.02 | N/A | CSF#1: 60<br>CSF #2: 69 | CSF #1: 58<br>CSF #2: 55 | CSF #1: 52<br>CSF #2: 59 | 57 | 55 |
| CSF protein, 15-45 mg/dL (mean ± SD) | 25.52 ± 10.46 | N/A | CSF#1: 79<br>CSF#2: 99 | CSF#1: 18<br>CSF #2: 24 | CSF#1: 28<br>CSF#2: 34 | 27 | 21 |
| CSF IgG, 0.9-4.8 mg/dL (mean ± SD) | 1.41 ± 1.12<br>**23 samples undetectable | N/A | CSF#1: 5.4<br>CSF #2: 6.9 |  | CSF#1: N/A<br>CSF#2: 3.0 | 1.5 |  |
| IgG index, 0.26-0.62 (mean ± SD) | 0.54 ± 0.08<br>**Unable to calculate for 23 samples | N/A | CSF#1: 0.54<br>CSF #2: 0.52 |  | CSF #1: 0.59<br>CSF #2: 0.55 | 0.58 | 0.48 |
| CSF albumin, 12-33 mg/dL (mean ± SD) | 11.72 ± 8.89 | N/A | CSF#1: 58<br>CSF #2: 61 | CSF#1: 9<br>CSF#2: 10 | CSF#1: N/A<br>CSF#2: 17 | 12 |  |
| Albumin quotient (3.2-9.0) |  | N/A | CSF#1: 13.55<br>CSF #2: 12.32 |  | CSF#1: N/A<br>CSF#2: 3.81 | 3.31 | 1.92 |

### Discovery of candidate autoantigens by PhIP-seq and IP-MS

PhIP-seq was performed to screen for autoantibodies in 84 CSF samples from 61 children with ASD, paired with 266 serum samples from these 61 children with ASD and 274 blood samples from 50 typically developing controls. In studies of autoantibody profiles from serum of healthy adults, it has been shown that each individual harbors a distinctive pattern of autoreactivities, termed the “autoreactome”^32^. The number of unique IgG autoantibodies in blood from healthy individuals is known to increase with age from infancy to adolescence, and thereafter plateaus^33^. We analyzed the CSF autoreactomes from ASD children, with a focus on the 6 children with anti-neural autoantibodies in CSF. While technical replicates showed high reproducibility (median Pearson’s *r* coefficient 0.79, interquartile range 0.75-0.88), the CSF autoreactivities from each case were distinct, with little similarity compared to other ASD cases (**Figure 2A**). Whereas prior PhIP-seq studies of peripheral blood from healthy adults showed longitudinally stable autoreactivities, the CSF autoreactomes from ASD cases with serial CSFs (Case 2, 3, and 4) demonstrated dynamic autoantibody profiles. These changes in CSF autoantibodies may parallel the acquisition of unique autoantibodies in blood during childhood to adolescence. Across the 6 cases with anti-neural immunoreactivity, we assessed for shared enrichment of antigenic targets to suggest an autoreactive “signature” (**Figure 2B, Table S1**).

**Figure 2.**
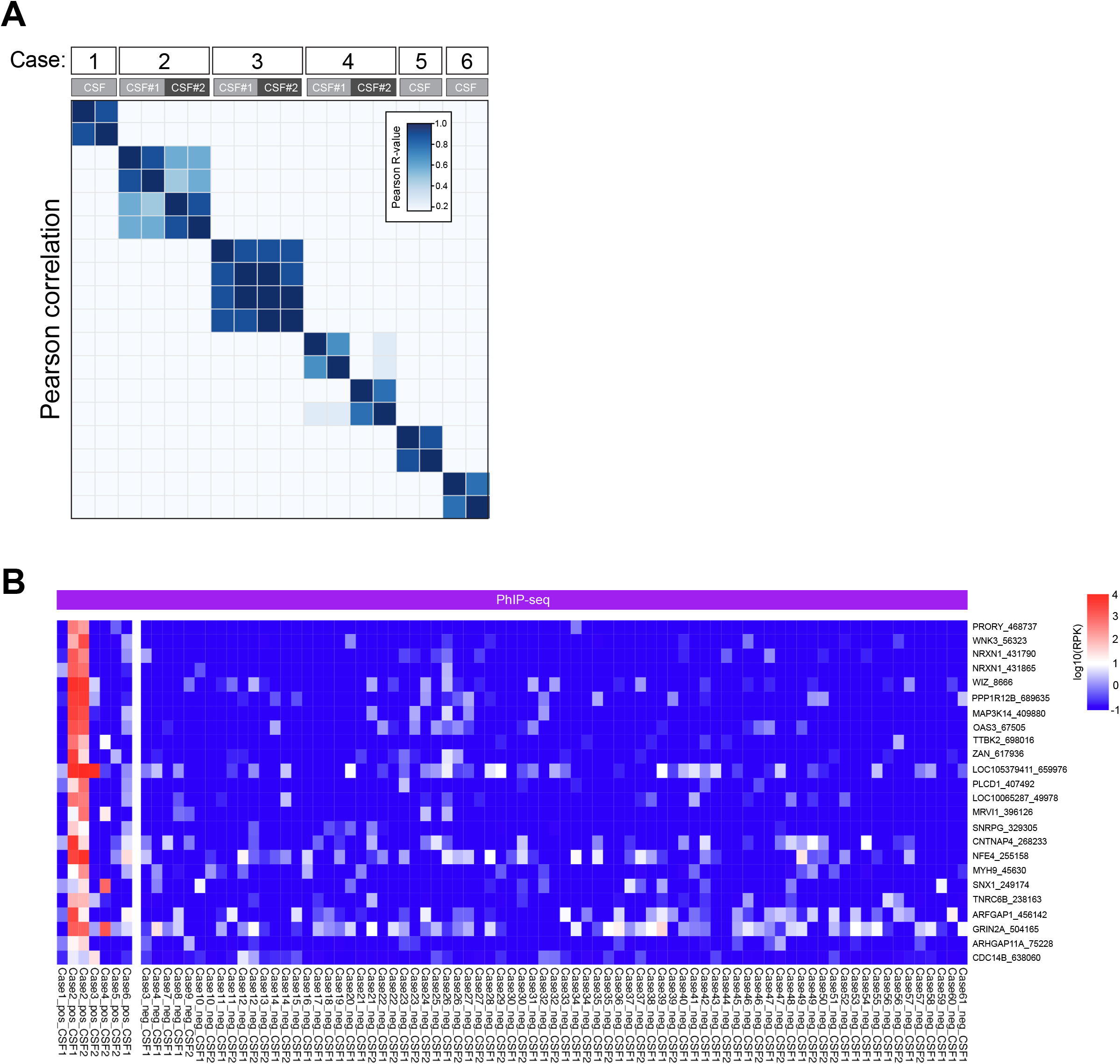
Characterization of autoreactivities in CSF from children with ASD. **(A)** Correlation matrix of Pearson *r* coefficients from PhIP-seq enrichments of CSF samples from 6 ASD cases that demonstrated anti-neural reactivities by tissue staining. Each row represents an individual technical replicate, and all CSF samples were included for each ASD case, even if not immunoreactive by tissue staining. **(B)** Heatmap of autoantigen enrichment from PhIP-seq of 84 ASD CSF samples, comparing 7 CSF samples from 6 ASD cases with positive anti-neural immunoreactivity (“Case#_pos”) versus 77 CSF samples from 55 ASD children with negative anti-neural immunoreactivity (“Case#_neg”). Data are represented as log10 bead normalized peptide reads. Each row represents a unique peptide fragment (“Gene”_”Peptide number”).

By this method, we identified several antigens with high-confidence genetic associations to ASD (**Table 2**) as defined by the SFARI Gene Database^34^.

**Table 2.** Strength of ASD genetic associations for top enriched CSF autoantigens shared in ≤ 2 cases with positive tissue staining.

| **Autoantigen** | **SFARI gene score\*** |
| --- | --- |
| NRXN1 | 1 |
| CNTNAP4 | 2 |
| MYH9 | 2 |
| TNRC6B | 2 |
| WNK3 | 2 |

\*SFARI Gene Score 1: High confidence
SFARI Gene Score 2: Strong candidate

Since we anticipate unique CSF autoantibodies in each case based on the distinct staining patterns and the differential clustering of autoreactivities by Pearson correlation, we also identified the antigenic enrichments for individual ASD cases compared to all negative-staining ASD CSF samples (**Figure 3, Table S1**). Candidate antigens from each patient were ranked based on PhIP-seq results, taking into account the normalized peptide counts, z-score, and a requirement that a peptide was enriched in both technical replicates. While PhIP-seq provides a human proteome-wide screen of autoreactivities, we leveraged IP-MS from mouse brain lysate to prioritize brain-specific antigens. Whole mouse brain lysate was immunoprecipitated with CSF from the 6 cases with anti-neural immunoreactivity on tissue staining along with mock bead-only controls. For each case, all proteins that met statistical criteria (log2FC over bead control > 1 and FDR-adjusted p value < 0.05) were considered present by IP (**Table S2**). We overlapped these IP-MS results with PhIP-seq peptide enrichments and further prioritized candidate brain-enriched antigens by evaluating their transcriptional expression in the human brain, as reported in the HPA Brain Resource^31^. From these curated lists of top candidate autoantigens per child, we noted multiple candidates were known to have a strong genetic association with ASD risk (**Table 3**). Notably, some of these targets have autoantibodies linked to described clinical syndromes, including anti-NRXN1α (subset of schizophrenia^35^), anti-SEZ6L2 (paraneoplastic cerebellar ataxia^36–38^), and anti-ANK3 (steroid-responsive meningoencephalitis^39^).

**Table 3.**
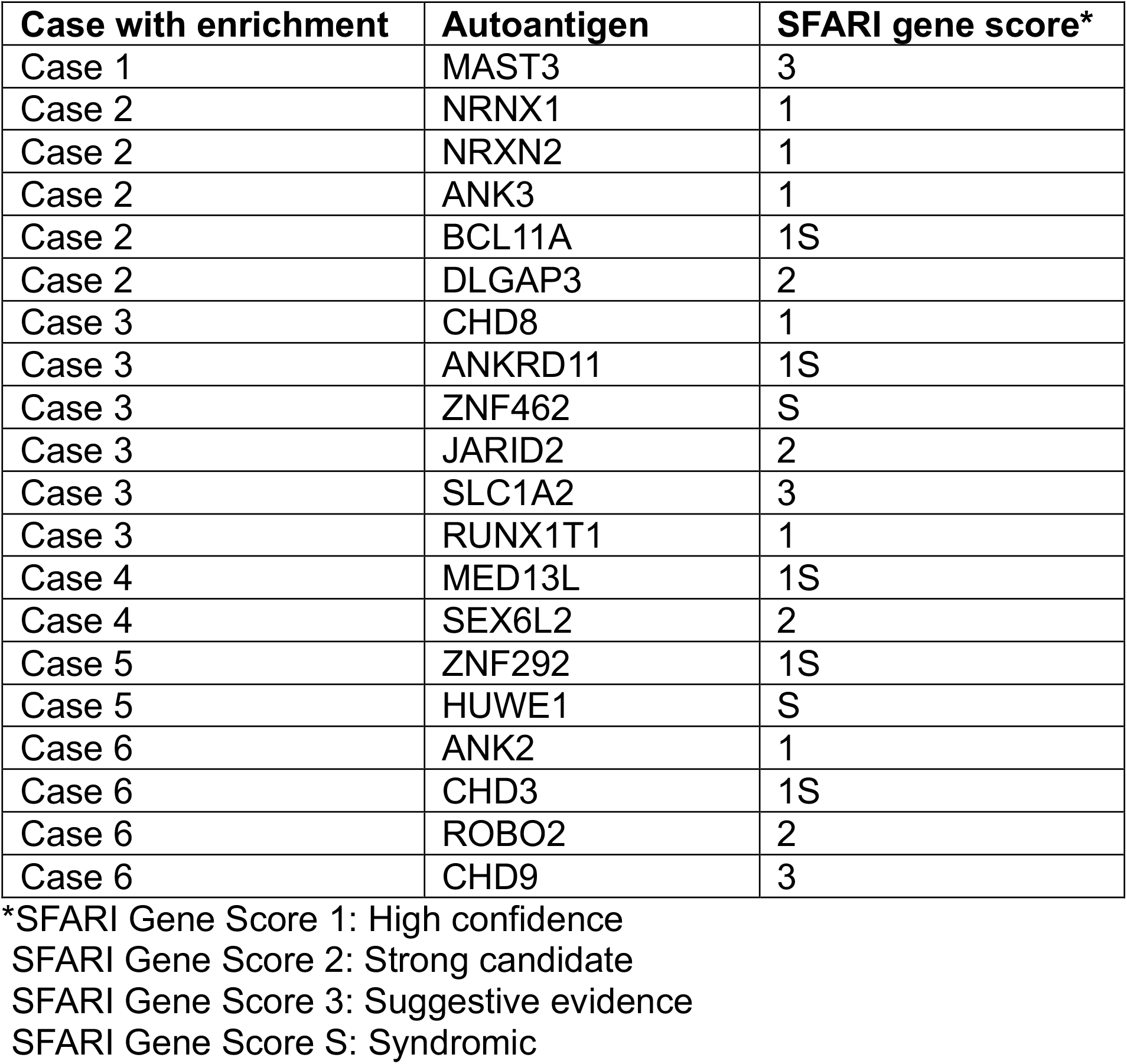
Strength of ASD genetic assocations for top CSF autoantigens enriched in individual cases with positive tissue staining.

**Figure 3.**
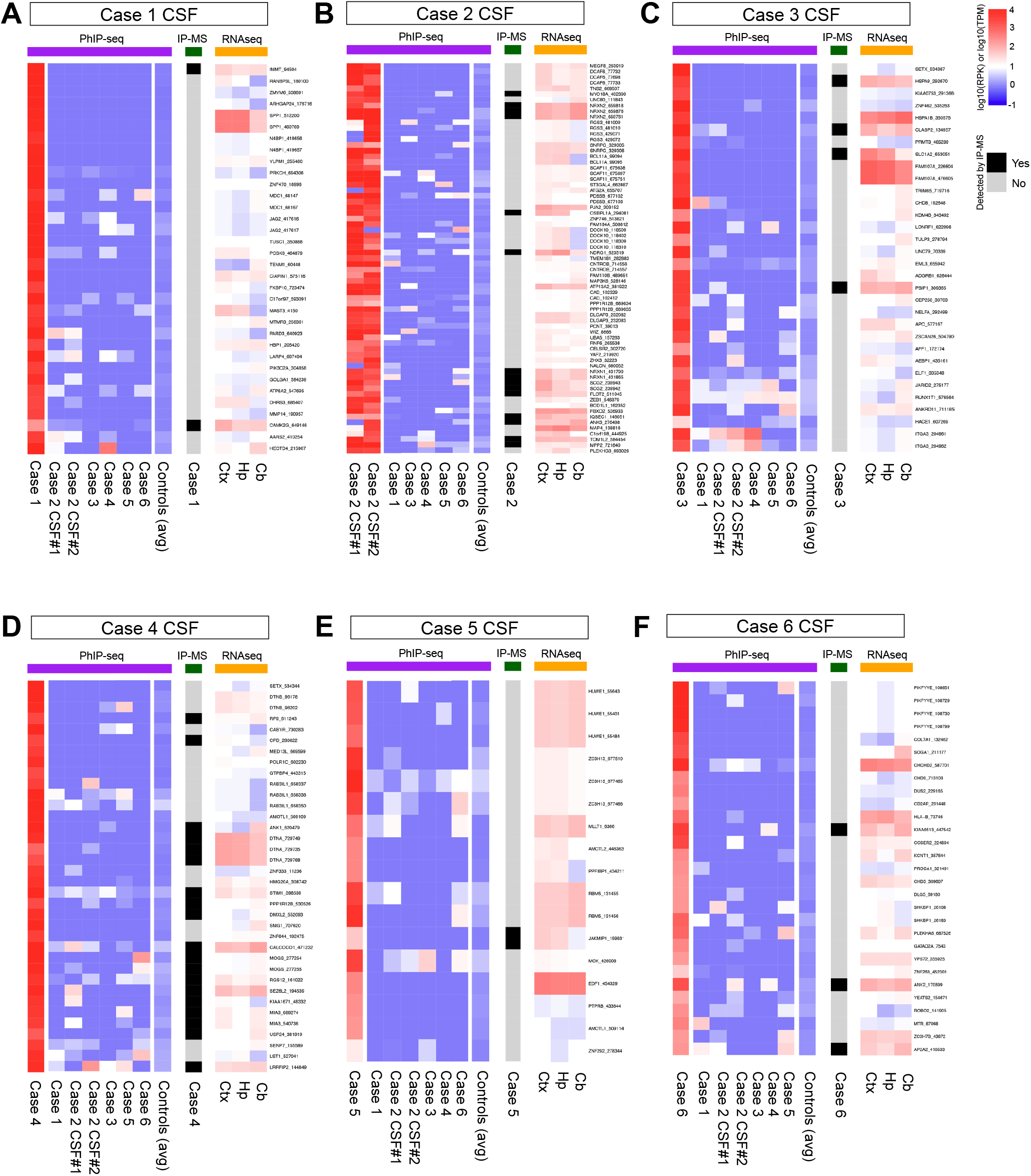
Profiling of autoantibody reactivities by PhIP-seq and IP-MS in ASD CSF with positive anti-neural immunoreactivity. Heatmap of autoantigen enrichments from PhIP-seq (purple bar), IP-MS (dark green bar), and RNA-seq from Human Protein Atlas Brain Resrouce (orange bar). Each heatmap displays PhIP-seq data as the enrichment of peptides in the index ASD case above the other ASD cases compared with the 77 CSF samples with negative anti-neural immunoreactivity (controls, averaged). IP-MS data is represented as the presence (black) or absence (grey) of the protein by this detection method. Data are represented as log10 normalized peptide reads for PhIP-seq and log10 transcripts per million (TPM) for RNA-seq. Each row represents a unique peptide fragment (“Gene”_”Peptide number”).

### Validation of candidate autoantibodies in Case 2

We focused our subsequent analyses to validate the candidate antigens targeted by Case 2’s CSF autoantibodies given the persistence of anti-neural antibodies in longitudinally collected CSF samples, which seem to display an expansive autoreactive repertoire that disproportionately drove the enrichments identified across the 6 ASD cases (**Figure 2B**). From comparing autoantigen candidates identified through PhIP-seq and IP-MS (**Figure 3B**), we filtered for proteins that were highly brain-enriched and have known functions in neurodevelopment. NRXN1 and NRXN2 were of particular interest given their known genetic association with ASD.

Multiple NRXN1 and NRXN2 peptide fragments were enriched by PhIP-seq, which mapped to the extracellular laminin G domain of the neurexin proteins’ beta isoforms (**Figure 4A**). Notably, the NRXN2 fragment was enriched nearly 30-fold higher than the NRXN1 fragment. Alignment of the NRXN1 and NRXN2 fragments by Clustal Omega^40^ revealed high sequence similarity between the peptide fragments (**Figure 4B**). PhIP-seq enrichments for anti-NRXN1β and anti-NRXN2β were elevated in the CSF compared to peripheral blood (**Figure 4C-D**). By overexpression of NRXN1-HA in HEK293T cells, presence of anti-NRXN1β in Case 2’s CSF failed to validate with immunocytochemistry and immunoblotting (**Figure 4E-F**). Next, we evaluated the presence of anti-NRXN2β by co-immunostaining mouse brain tissue with Case 2’s CSF and a commercially available anti-NRXN2 antibody and identified colocalization of immunofluorescent signal in CA3 region of the hippocampus (**Figure 4G**). Since we identified elevated anti-NRXN2β in blood samples from one control (Control 24) and in Case 2 (**Figure 4D**), we performed a cell-based assay with overexpression of NRXN2-FLAG in HEK293T cells, which did not demonstrate colocalization of peripheral blood autoantibodies in either Case 2 or Control 24 with NRXN2-FLAG (**Figure 4H**). Additional confirmation of anti-NRXN2 in CSF through orthogonal assays, including cell-based assays and immunoblotting, will provide additional evidence to motivate further testing of functional impact and disease relevance of anti-NRXN2 autoantibodies.

**Figure 4.**
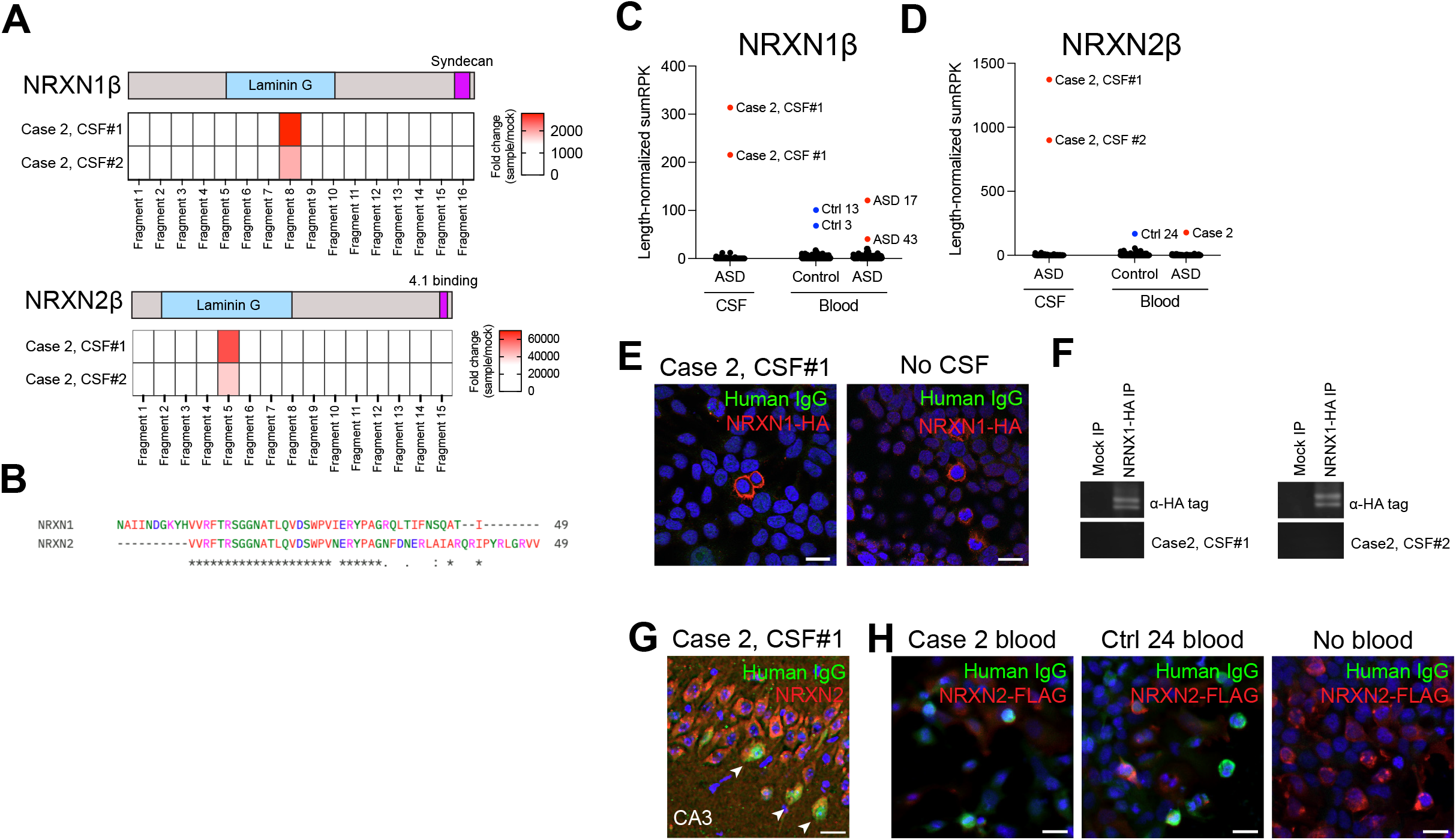
Validating anti-neurexin autoantibodies in ASD CSF. **(A)** Epitope map of NRXN1β and NRXN2β peptides enriched by Case 2 CSF antibodies in PhIP-seq. Data are shown as PhIP-seq normalized reads (fold change over mock bead control) per peptide fragment. **(B)** Alignment of NRXN1β Fragment 8 and NRXN2β fragment 5 shows sequence similarities. **(C)** Enrichments in NRXN1β and **(D)** NRXN2β proteins across CSF and blood samples from ASD and typically developing children (Control, Ctrl) screened by PhIP-Seq. Enrichments are represented as length-normalized reads (total reads for the protein/number of peptides that map to the protein in the PhIP-seq library). **(E)** No immunoreactivity detected from Case 2 CSF to HEK293T cells overexpressing HA-tagged NRXN1 compared to no CSF control. CSF dilution 1:10. Scalebars, 20μm. **(F)** Immunoblot of HEK293T cells lysates overexpressing HA-tagged NRXN1 probed with Case 2 CSF#1 and CSF#2 samples. CSF dilution 1:10. **(G)** Immunostaining of mouse brain tissue with Case 2 CSF at 1:4 dilution (green) and anti-NRXN1 (red) shows colocalization in hippocampal neurons in CA3 region. Scalebars, 20μm. **(H)** No immunoreactivity detected from Case 2 and Ctrl 24 blood antibodies at 1:200 dilution to HEK293T cells overexpressing FLAG-tagged NRXN2 compared to no CSF control. CSF dilution 1:10. Scalebars, 20μm.

## DISCUSSION

ASD is a heterogeneous condition affecting a significant number of children worldwide, where more than 80% of cases are without a known cause. While genetic and immune risk factors have been identified, the neurobiological mechanisms leading to the social and behavioral deficits in ASD remain elusive. We profiled CSF and blood autoantibodies from children with ASD and identified that among 6 children with anti-neural CSF autoreactivity, brain tissue immunostaining revealed distinct anatomical and subcellular patterns. These case-specific immunostaining patterns suggest that each child harbors a unique repertoire of CSF autoantibodies. How these anti-neural antibodies do or do not relate to ASD remains to be determined. Using complementary screening approaches (PhIP-seq and IP-MS), we curated a list of top candidate autoantigens per child. Notably, multiple of these top candidates have known high-confidence associations with ASD (ANK2/3, CHD8, NRXN1/2, RUNX1T1) along with known genetic syndromes in which ASD is a common feature (ANKRD11, BCL11A, CHD3, MED13L, ZNF292). While confirmation of these autoantibody reactivities remains essential, the convergence of these autoantigens with genetic risk suggests shared neurobiological pathways that underlie the pathophysiology of ASD.

Functionally, these top candidate autoantigens can be classified into several molecular pathways: 1) Synaptic connectivity (NRXN1/2), 2) Neuronal scaffolding (ANK2/3), and 3) Transcriptional and chromatin regulation that may govern neurodevelopmental gene programs (BCL11A, CHD3, CHD8, MED13L, RUNX1T1, ZNF292). Autoantibodies against cell surface or secreted proteins could have a direct pathogenic effect in perturbing synaptic function and plasticity by function-blocking properties of antibody binding versus recruitment of microglia, whereas autoantibodies against intracellular components may instead be evidence of cellular injury and potentially serve as ASD biomarkers rather directly have a pathogenic role. Therefore, prioritizing validation of cell surface and extracellularly expressed candidate autoantigens is crucial to evaluating whether the associated autoantibodies have a role in the pathophysiology of ASD.

Neurexins, which were identified in our autoantibody screen, are a prime example of a cell surface protein with known functions in synaptic structure and function. NRXN2β is a single-pass membrane-bound protein with an extracellular domain. The laminin G domain of neurexins undergoes calcium-dependent interactions with postsynaptic binding partners, including neuroligins^41^, dystroglycans, cerebellins, and Leucin-Rich Repeat Transmembrane proteins (LRRTMs)^42^. Interactions with multiple binding partners allow neurexins to function as synaptic “hub” molecules to shape synaptic connectivity and function^43^. Truncating mutations in *NRXN2*, leading to loss of laminin G domains and subsequent failure to associate with postsynaptic neuroligin and LRRTM binding partners, has been identified in patients with ASD^44^. By PhIP-seq, we detected enrichment of a peptide within the NRXN2β laminin G domain. As of yet, we have been unable to orthogonally validate this as a bona fide autoantigen in this case, but its potential relevance to ASD is intriguing.

By comparing CSF and blood autoantibody profiles, we identified CNS-specific immune processes that were not mirrored in the peripheral blood. Moreover, identification of CNS-enriched candidate autoantibodies may reveal chronic neurobiological pathway disruptions that were not captured in prior investigations of the cytokine and chemokine profiles that were more reflective of acute immune states. Together, our findings broaden the lens on autoimmunity in ASD by identifying candidate autoantibodies targeting members of synaptic, scaffolding, and transcriptional pathways.

## Limitations

This study lacks comparison data for CSF from typically developing pediatric controls, as there are ethical constraints to performing LPs on healthy children. With complementary high-throughput screening methods, we have identified candidate autoantibodies in CSF from children with ASD, although whether they impact neurodevelopment remains to be determined. While there is substantial evidence connecting autoimmunity and ASD, it is unclear whether immune dysfunction is causal or a sequela of the disorder. We prioritized our downstream analyses on ASD cases for whom anti-neural immunofluorescence on rodent brain tissues was detected. However, we recognize that this approach may miss autoreactivities against neural antigens that lack a rodent ortholog. Lastly, PhIP-seq has poor sensitivity for conformational and post-translationally modified epitopes. Nevertheless, we demonstrate the potential for unbiased assays in screening for autoantigens associated with neurodevelopmental disorders.

## Data Availability

All data produced in the present study are available upon reasonable request to the authors.

## ACKNOWLEDGEMENTS

This work was supported by R01MH122471 (PIs SJP and MRW). MPL is a Fellow in the Pediatric Scientist Development Program. This project was supported by the American Academy of Pediatrics (AAP) and the American Pediatric Society (APS). CMB is supported by NIMH IRP ZIA MH002982. This research was supported in part by the Intramural Research Program of the National Institutes of Health (NIH). The contributions of the NIH author(s) are considered Works of the United States Government. The findings and conclusions presented in this paper are those of the author(s) and do not necessarily reflect the views of the NIH or the U.S. Department of Health and Human Services.

